# Treatment of Severe COVID-19 with Convalescent Plasma in the Bronx, NYC

**DOI:** 10.1101/2020.12.02.20242909

**Authors:** Hyun ah Yoon, Rachel Bartash, Inessa Gendlina, Johanna Rivera, Antonio Nakouzi, Robert H. Bortz, Ariel S. Wirchnianski, Monika Paroder, Karen Fehn, Leana Serrano-Rahman, Rachelle Babb, Uzma N. Sarwar, Denise Haslwanter, Ethan Laudermilch, Catalina Florez, M. Eugenia Dieterle, Rohit K. Jangra, J. Maximilian Fels, Karen Tong, Margarette C. Mariano, Olivia Vergnolle, George I. Georgiev, Natalia G. Herrera, Ryan J. Malonis, Jose A. Quiroz, Nicholas C. Morano, Gregory J. Krause, Joseph M. Sweeney, Kelsie Cowman, Stephanie Allen, Jayabhargav Annam, Ariella Applebaum, Daniel Barboto, Ahmed Khokhar, Brianna J. Lally, Audrey Lee, Max Lee, Avinash Malaviya, Reise Sample, Xiuyi A. Yang, Yang Li, Rafael Ruiz, Raja Thota, Jason Barnhill, Doctor Y. Goldstein, Joan Uehlinger, Scott J. Garforth, Steven C. Almo, Jonathan R. Lai, Morayma Reyes Gil, Amy S. Fox, Kartik Chandran, Tao Wang, Johanna P. Daily, Liise-anne Pirofski

## Abstract

Convalescent plasma with severe acute respiratory disease coronavirus 2 (SARS-CoV-2) antibodies (CCP) may hold promise as treatment for Coronavirus Disease 2019 (COVID-19). We compared the mortality and clinical outcome of patients with COVID-19 who received 200mL of CCP with a Spike protein IgG titer ≥1:2,430 (median 1:47,385) within 72 hours of admission to propensity score-matched controls cared for at a medical center in the Bronx, between April 13 to May 4, 2020. Matching criteria for controls were age, sex, body mass index, race, ethnicity, comorbidities, week of admission, oxygen requirement, D-dimer, lymphocyte counts, corticosteroids, and anticoagulation use. There was no difference in mortality or oxygenation between CCP recipients and controls at day 28. When stratified by age, compared to matched controls, CCP recipients <65 years had 4-fold lower mortality and 4-fold lower deterioration in oxygenation or mortality at day 28. For CCP recipients, pre-transfusion Spike protein IgG, IgM and IgA titers were associated with mortality at day 28 in univariate analyses. No adverse effects of CCP were observed. Our results suggest CCP may be beneficial for hospitalized patients <65 years, but data from controlled trials is needed to validate this finding and establish the effect of ageing on CCP efficacy.

## Introduction

Severe acute respiratory syndrome coronavirus-2 (SARS-CoV-2) (1), a highly transmissible enveloped positive-strand RNA virus (2), is the causative agent of the Coronavirus disease 2019 (COVID-19) pandemic (3). The first case was reported in December 2019, and by November 2020 more than 61 million infections were reported worldwide, with one fifth of the cases and deaths occurring in the United States (U.S.) (4). In April 2020, New York City (NYC), especially the borough of the Bronx, was an early epicenter of the COVID-19 pandemic in the U.S. (5, 6). Since then, an antiviral that reduced duration of illness (7), Remdesivir, received FDA approval on October 22, 2020, and corticosteroids, which reduced mortality in severely ill patients in a large randomized clinical trial and prospective meta-analysis (8, 9), have become standard of care. As of November 2020, there is no approved therapy for COVID-19 that reduces mortality of hospitalized patients with respiratory manifestations of severe or life-threatening disease.

Convalescent plasma (CP) obtained from recovered persons was deployed for previous respiratory pandemics, including 1918 and 2009 influenza and Severe Acute Respiratory Syndrome (SARS) (10-13). Given the lack of established therapies for COVID-19, CP containing SARS-CoV-2 antibodies (CCP) was proposed as a therapeutic option early in the pandemic (14). As of November 2020, it has shown a possible benefit in multiple studies. In early case series, CCP-treated patients exhibited viral clearance and reductions in inflammatory markers (15-19). Observational studies comparing CCP-treated patients to retrospective controls showed a reduction in mortality in non-intubated patients and/or those transfused within 72 hours of hospitalization with high titer CCP (20-24). Analysis of a subset of more than 3,000 CCP recipients in an open label study found a dose-response whereby non-intubated patients who received high titer CCP had lower mortality than those who received low titer CCP (22). Several randomized controlled trials (RCT) have not shown a benefit of CCP, but were limited by premature termination due to a lack of patients to recruit (25, 26). One trial found CCP had an antiviral effect, but there was no effect on mortality (27), another found a reduction in mortality, albeit with a very small sample size (28), and another was terminated due to the presence of neutralizing antibodies in CCP recipients at the time of transfusion, despite being on track to meet the primary endpoint (29). A recent double-blind, placebo controlled, multicenter RCT did not show an effect of CCP on mortality (30). Evidence of safety and possible benefit led the FDA to issue an emergency use authorization (EUA) for CCP use in hospitalized patients with COVID-19 on August 23, 2020.

We treated 103 patients at Montefiore Medical Center (MMC), a 1,491-bed tertiary medical center in the Bronx, New York with serious or life-threatening COVID-19 with CCP between April 13 and May 4, 2020 and conducted a propensity score matched study. Herein, we report mortality and clinical and laboratory findings of CCP recipients compared to matched controls.

## Results

### Baseline characteristics of CCP recipients and retrospective controls

One hundred and three (103) patients were enrolled in the Mayo Clinic expanded access protocol (EAP) (31) and treated with one 200 mL unit of CCP within 72 hours of hospital admission. Clinical status and mortality on day 28 post-transfusion was compared to retrospective propensity score matched controls identified by querying the electronic medical record (Figure 1). Analysis included 90 CCP recipients and 258 controls after exclusion of twelve patients who did not meet the eligibility criteria because of mechanical ventilation for > 24 hours or CCP transfusion > 72 hours after admission, and one patient with missing data (Figure 1). Compared to controls, CCP recipients were younger (median 66 vs 72 years, *P* = 0.002), had higher BMI (28 vs 27 kg/m^2^, *P* = 0.05), and lower rates of congestive heart failure (18 vs 29%, *P* = 0.03), and chronic kidney disease (29 vs 41%, *P* = 0.03). At baseline, a lower proportion of CCP recipients were on low-flow oxygen support (68 vs 81%, *P* < 0.0001) and a higher proportion required mechanical ventilation (20 vs 6%, *P* < 0.0001), had lower baseline lymphocyte counts (0.8 vs 1.0 x 10^9^/L, *P* = 0.001), and received systemic corticosteroids (93 vs 63%, *P* < 0.0001) (Supplemental Table 1). After propensity score matching, 73 CCP recipients and 73 control patients were well balanced for all matching variables except in a subgroup analysis stratified by age; CCP recipients ≥ 65 years had lower lymphocyte counts than controls (median, 0.8 vs 1.0 x10^9^/L) (Table 1, Supplemental Figure 1).

**Table 1.** Baseline characteristics and outcomes in CCP recipients transfused by admission day 3 (n = 73) and propensity score matched controls (n = 73).

|  | All patients |  |  | < 65 yrs |  |  | ≥ 65 yrs |  |  |
| --- | --- | --- | --- | --- | --- | --- | --- | --- | --- |
| Characteristic | Case<br>(n=73; 64<br>not on MV,<br>9 on MV) | Controls<br>(n=73; 64<br>not on MV,<br>9 on MV) | P value | Case (n=34;<br>28 not on<br>MV, 6 on<br>MV) | Controls<br>(n=34; 28<br>not on MV,<br>6 on MV) | P value | Case (n=39;<br>36 not on<br>MV, 3 on<br>MV) | Controls<br>(n=39; 36<br>not on MV,<br>3 on MV) | P value |
| Median age<br>(IQR) - yr | 67 (55-75) | 66 (56-77) | 0.80 | 55 (50-57) | 55 (49-59) | 0.84 | 75 (69-82) | 76 (71-81) | 0.63 |
| Not on MV | 68 (55-77) | 67 (56-77.5) | 0.87 | 54.5 (48.5-59) | 55.5 (48.5-60) | 0.86 | 75 (70-82) | 76.5 (71-82) | 0.80 |
| On MV | 56 (55-66) | 59 (53-72) | 0.93 | 55.5 (50-56) | 53.5 (49-59) | 0.87 | 69 (66-78) | 75 (72-78) | 0.38 |
| **Age category – no. (%), yr** |  |  |  |  |  |  |  |  |  |
| <45 | 7 (9.6) | 6 (8.2) | 0.97 | 7 (20.6) | 6 (17.7) | 0.76 | 0 | 0 |  |
| 45 to <65 | 27 (36.9) | 28 (38.4) |  | 27 (79.4) | 28 (82.4) |  | 0 | 0 |  |
| 65 to <75 | 19 (26.0) | 17 (23.3) |  | 0 | 0 |  | 19 (48.7) | 17 (43.6) | 0.82 |
| 75 to <85 | 14 (19.2) | 14 (19.2) |  | 0 | 0 |  | 14 (35.9) | 14 (35.9) |  |
| ≥85 | 6 (8.2) | 8 (10.9) |  | 0 | 0 |  | 6 (15.4) | 8 (20.5) |  |
| **Sex, BMI** |  |  |  |  |  |  |  |  |  |
| Male – no. (%) | 41 (56.2) | 47 (64.4) | 0.31 | 21 (61.8) | 22 (64.7) | 0.80 | 20 (51.3) | 25 (64.1) | 0.25 |
| Body-mass index, median (IQR)<sup>A</sup> | 28.3 (23.7-33.5) | 27.4 (23.2-31.6) | 0.32 | 29.6 (27.6-34.7) | 28.1 (24.8-34.3) | 0.52 | 26.9 (21.9-32.8) | 25.8 (22.9-28.5) | 0.58 |
| **Ethnicity<sup>B</sup>– no. (%)** |  |  |  |  |  |  |  |  |  |
| Hispanic | 38 (52.1) | 36 (49.3) | 0.45 | 16 (47.1) | 18 (52.9) | 0.39 | 22 (56.4) | 18 (46.2) | 0.68 |
| Non-Hispanic | 24 (32.9) | 30 (41.1) |  | 11 (32.4) | 13 (38.2) |  | 13 (33.3) | 17 (43.6) |  |
| Unknown | 11 (15.1) | 7 (9.6) |  | 7 (20.6) | 3 (8.8) |  | 4 (10.3) | 4 (10.3) |  |
| **Race<sup>B</sup>– no. (%)** |  |  |  |  |  |  |  |  |  |
| White | 6 (8.2) | 6 (8.2) | 0.93 | 3 (8.8) | 4 (11.8) | 0.56 | 3 (7.7) | 2 (5.1) | 0.79 |
| African American | 19 (26.0) | 17 (23.3) |  | 12 (35.3) | 8 (23.5) |  | 7 (17.9) | 9 (23.1) |  |
| Others/Unknown | 48 (65.8) | 50 (68.5) |  | 19 (55.9) | 22 (64.7) |  | 29 (74.4) | 28 (71.8) |  |
| **Coexisting disorder – no. (%)** |  |  |  |  |  |  |  |  |  |
| Hyperlipidemia | 42 (57.5) | 44 (60.3) | 0.74 | 17 (50.0) | 15 (44.1) | 0.63 | 25 (64.1) | 29 (74.4) | 0.33 |
| Hypertension | 60 (82.2) | 62 (84.9) | 0.66 | 24 (70.6) | 27 (79.4) | 0.40 | 36 (92.3) | 35 (89.7) | 0.69 |
| Coronary artery disease | 10 (13.7) | 13 (17.8) | 0.49 | 2 (5.9) | 4 (11.8) | 0.67 | 8 (20.5) | 9 (23.1) | 0.78 |
| Congestive heart failure | 15 (20.6) | 19 (26.9) | 0.43 | 4 (11.8) | 5 (14.7) | 0.72 | 11 (28.2) | 14 (35.9) | 0.47 |
| Chronic pulmonary disease | 26 (35.6) | 22 (30.1) | 0.48 | 10 (29.4) | 7 (20.6) | 0.40 | 16 (41.0) | 15 (38.5) | 0.82 |
| Chronic kidney disease | 24 (32.9) | 31 (42.5) | 0.23 | 8 (23.5) | 9 (26.5) | 0.78 | 16 (41.0) | 22 (56.4) | 0.17 |
| Obesity (BMI ≥ 30) | 29 (39.7) | 22 (30.1) | 0.22 | 16 (47.1) | 15 (44.1) | 0.81 | 13 (33.3) | 7 (17.9) | 0.12 |
| Morbid obesity (BMI ≥ 35) | 14 (19.2) | 10 (13.7) | 0.37 | 7 (20.6) | 7 (20.6) | 1.00 | 7 (17.9) | 3 (7.7) | 0.18 |
| Diabetes | 33 (45.2) | 34 (46.6) | 0.87 | 11 (32.4) | 11 (32.4) | 1.00 | 22 (56.4) | 23 (58.9) | 0.82 |
| **Pharmacologic interventions – no. (%)** |  |  |  |  |  |  |  |  |  |
| Corticosteroids | 67 (91.8) | 64 (87.7) | 0.41 | 29 (85.3) | 30 (88.2) | 0.72 | 38 (97.4) | 34 (87.2) | 0.09 |
| Therapeutic anticoagulation | 56 (76.7) | 47 (64.4) | 0.10 | 18 (52.9) | 26 (76.5) | **0.04** | 29 (74.4) | 30 (76.9) | 0.79 |

| Oxygen on the day of transfusion – no. (%) |  |  |  |  |  |  |  |  |  |
| --- | --- | --- | --- | --- | --- | --- | --- | --- | --- |
| Low-flow oxygen, 5L - 15L | 53 (72.6) | 53 (72.6) | 1.00 | 23 (67.7) | 23 (67.7) | 1.00 | 30 (76.9) | 30 (76.9) | 1.00 |
| High-flow oxygen or NIV | 11 (15.1) | 11 (15.1) |  | 5 (14.7) | 5 (14.7) |  | 6 (15.4) | 6 (15.4) |  |
| MV | 9 (12.3) | 9 (12.3) |  | 6 (17.7) | 7 (17.7) |  | 3 (7.7) | 5 (7.7) |  |
| Laboratory values on the day of transfusion, median (IQR) |  |  |  |  |  |  |  |  |  |
| Lymphocyte count, x10<sup>9</sup>/L | 0.8 (0.5-1.2) | 0.9 (0.7-1.2) | 0.97 | 0.9 (0.7-1.2) | 0.9 (0.7-1.2) | 0.44 | 0.8 (0.5-1.0) | 1.0 (0.6-1.2) | **0.03** |
| Not on MV | 0.8 (0.6-1.1) | 0.9 (0.7-1.2) | 0.12 | 0.9 (0.7-1.2) | 0.9 (0.7-1.2) | 0.90 | 0.8 (0.5-1.0) | 1.0 (0.6-1.4) | **0.04** |
| On MV | 1.2 (0.5-3.9) | 1.2 (0.9-1.2) | 0.72 | 1.3 (0.8-3.9) | 1.1 (0.9-1.3) | 0.47 | 0.5 (0.2-8.8) | 1.2 (0.8-1.2) | 0.51 |
| D-dimer, µg/ml | 2.8 (1.2-8.1) | 2.9 (1.4-6.4) | 0.83 | 1.5 (0.7-7.7) | 2.3 (1.3-5.1) | 0.27 | 3.7 (2.1-10.9) | 3.2 (1.9-7.4) | 0.41 |
| Not on MV | 2.8 (1.2-7.5) | 3.1 (1.4-7.1) | 0.51 | 1.2 (0.6-3.0) | 2.3 (1.2-6.3) | 0.09 | 3.8 (2.1-9.9) | 3.3 (2.1-7.9) | 0.43 |
| On MV | 7.7 (2.2-9.3) | 2.1 (1.6-3.6) | 0.20 | 7.9 (2.3-9.3) | 2.2 (1.6-3.6) | 0.15 | 2.2 (1.0-17.1) | 2.1 (1.2-4.3) | 0.83 |
| C-reactive protein, mg/dL | 17.4 (11.1-27.9) | 17.8 (8.3-28.5) | 0.68 | 16.2 (10.6-24.2) | 17 (7.1-27.6) | 0.97 | 18.8 (14.8-28.8) | 18.2 (8.5-28.7) | 0.52 |
| Not on MV | 18.1 (11-28.2) | 18.2 (8.1-28.9) | 0.86 | 16.2 (9.6-23.3) | 14.5 (6.8-29.2) | 0.92 | 19.5 (14.9-28.9) | 18.5 (8.8-28.8) | 0.68 |
| On MV | 14.9 (11.2-23.1) | 12 (9.1-20.1) | 0.56 | 16.5 (11.5-28.0) | 18.6 (9.1-25) | 0.87 | 11.9 (8.7-15) | 9.6 (4.3-12.0) | 0.56 |
| **Mortality Outcomes – no. (%)** |  |  |  |  |  |  |  |  |  |
| Day 28 mortality | 23 (31.5) | 28 (38.4) | 0.39 | 3 (8.8) | 10 (29.4) | **0.03** | 20 (52.6) | 17 (45.9) | 0.56 |
| Not on MV | 21 (32.8) | 24 (37.5) | 0.58 | 2 (7.1) | 8 (28.6) | **0.04** | 19 (52.8) | 16 (44.4) | 0.48 |
| On MV | 2 (22.2) | 4 (44.4) | 0.32 | 1 (14.2) | 3 (42.8) | 0.23 | 1 (33.3) | 2 (66.7) | 0.41 |
| **Day 28 clinical status – no. (%)** |  |  |  |  |  |  |  |  |  |
| Stable/better | 47 (64.4) | 42 (57.5) | 0.39 | 30 (88.2) | 22 (64.7) | **0.02** | 17 (43.6) | 20 (51.3) | 0.49 |
| Worse/dead | 26 (35.6) | 31 (42.5) |  | 4 (11.8) | 12 (35.3) |  | 22 (56.4) | 19 (48.7) |  |

Abbreviations: BMI, body mass index; IQR, interquartile range; MV, mechanical ventilation; NIV, noninvasive ventilation
A. Body-mass index is the weight in kilograms divided by the square of the height in meters.
B. Information on race and ethnic group was obtained from entries in the medical record, as reported by the patients.

**Figure 1.**
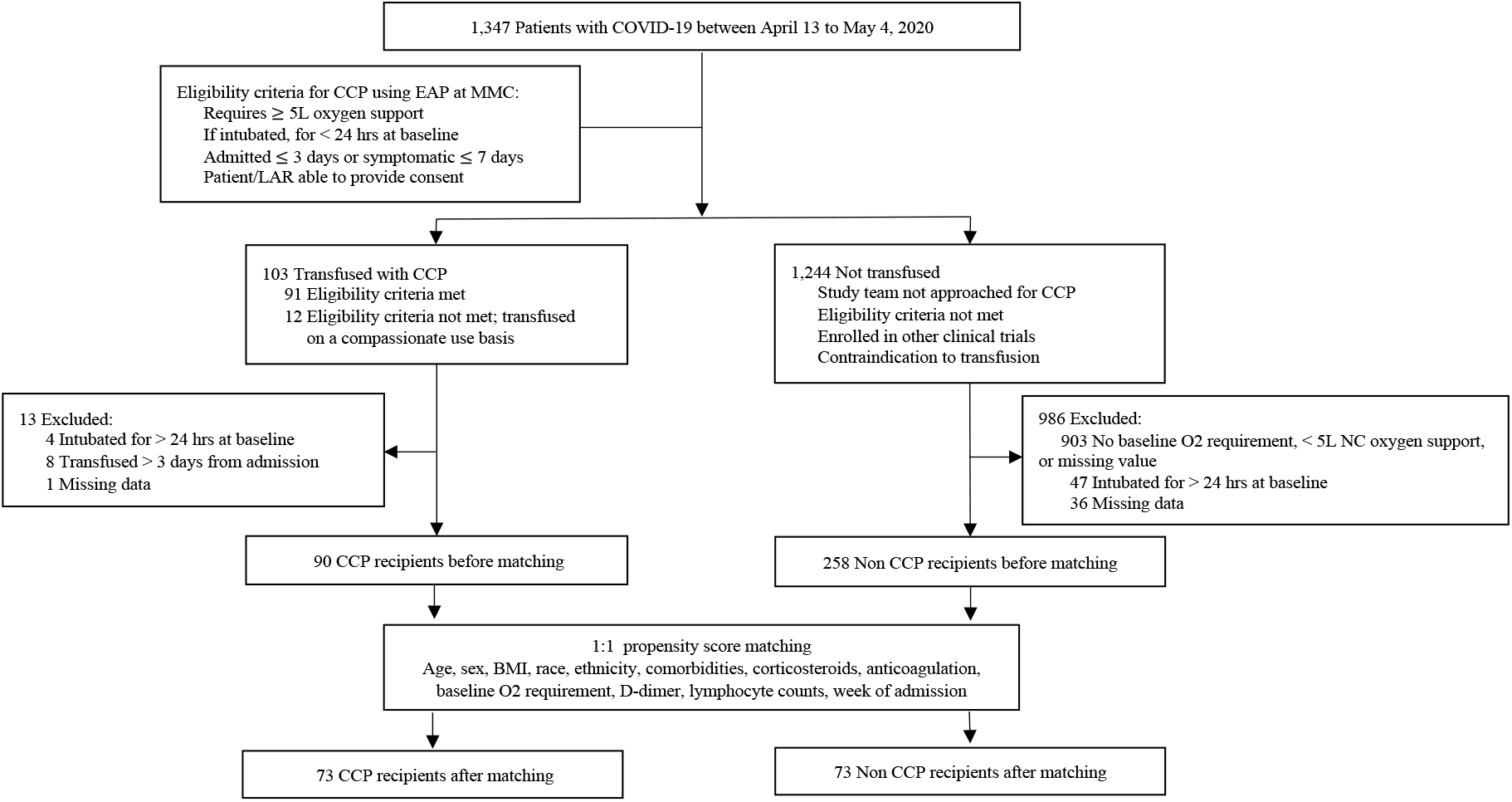
Enrollment of Study Patients and Distribution of Study Cohorts. Study baseline was defined as time of CCP transfusion for CCP recipients and admission day 2 for non CCP recipients. COVID-19, coronavirus disease 2019; CCP, COVID-19 convalescent plasma; EAP, expanded access protocol; LAR, legally authorized representative; MMC, Montefiore Medical Center; NC, nasal cannula.

### Convalescent plasma SARS-CoV-2 spike protein and neutralizing antibody (NAb) titers

Of the 200 mL units of CCP administered in this study, 95 of 103 were obtained from 46 persons who donated CP at MMC in April 2020. SARS-CoV-2 spike protein IgG endpoint titers were measured in CCP obtained from these donors using the MMC in-house research full-length spike protein ELISA (32). Median IgG, IgM and IgA titers were, respectively, 1:47,385 (interquartile range [IQR], 21,870 – 65,610; n = 46), 1:810 (IQR, 810 – 2,430; n = 43) and 1:90 (IQR, 90 – 270; n = 43). Median NAb titer by pseudovirus neutralization assay was 1:938 (IQR, 407 – 2,784; n = 42). There was a direct correlation between NAb titer and spike protein IgG (Spearman *r* = 0.78, *P* < 0.0001), IgM (*r* = 0.58, *P* < 0.0001) and weakly with IgA (*r* = 0.29, *P* = 0.05) titers (Supplemental Figure 2). For the eight patients who did not receive MMC donor CCP, 7 received units that tested as ‘reactive’ by the New York State Department of Health (NYSDOH) Wadsworth Center’s SARS-CoV-2 Microsphere Immunoassay for anti-Nucleocapsid antibody detection (33), and one received CCP with a spike protein IgG titer of 1:320 measured by the in-house spike protein ELISA assay at Mount Sinai Hospital (34).

### Comparison of clinical outcomes of CCP recipients and controls

There was no difference in mortality between 73 CCP recipients and 73 propensity score matched controls by day 28 (*P* = 0.47, Kaplan-Meier log rank test) (Figure 2). To account for the potential interaction between age and CCP treatment (*P* = 0.11, interaction term), analysis was stratified by age. Compared to matched controls, CCP recipients < 65 years had lower mortality by day 28 (*P* = 0.04, Kaplan-Meier log rank test), whereas the mortality of CCP recipients and matched controls ≥ 65 years did not differ significantly (*P* = 0.61, Kaplan-Meier log rank test) (Figure 2). There was no difference in mortality between groups when CCP recipients and controls were stratified by baseline oxygen requirement (Supplemental Figure 3).

**Figure 2.**
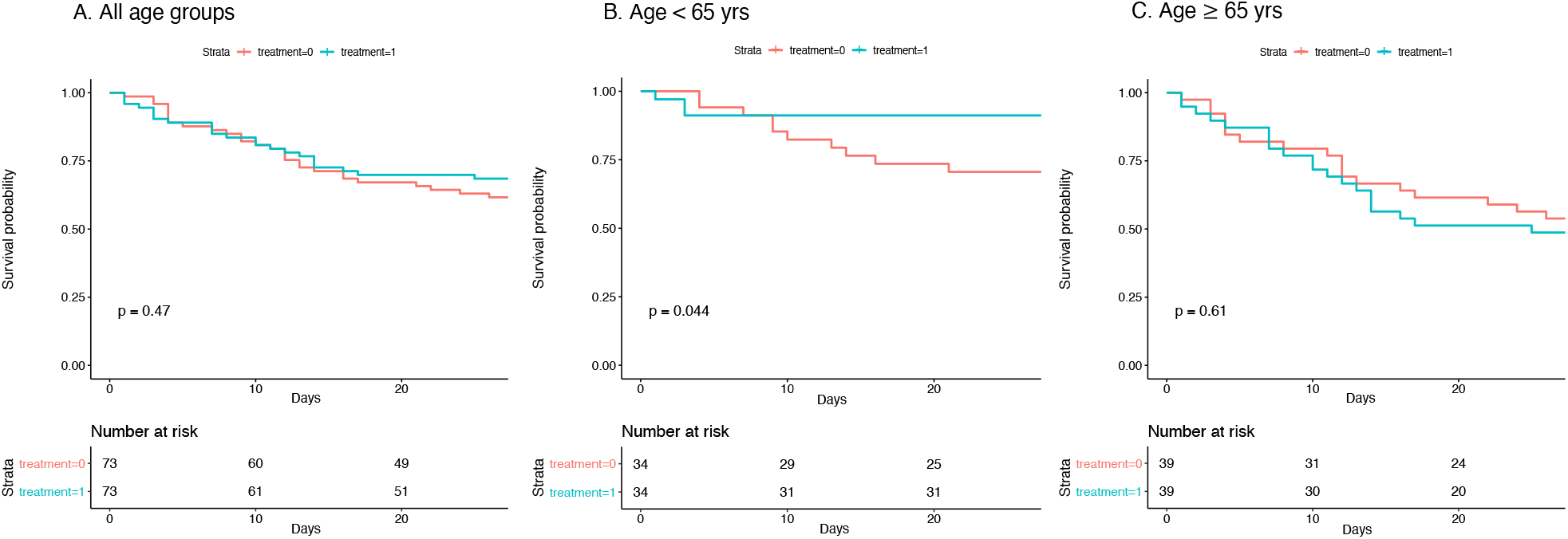
Kaplan–Meier Plot of the Probability of Survival from time of transfusion to day 28 in CCP recipients (n = 73) vs matched controls (n = 73). A. All age groups. B. Age < 65 years. C. Age ≥ 65 years.

There was no significant difference between CCP recipients and matched controls in all-cause mortality at 28 days (31.5 vs 38.4%; odds ratio [OR], 0.74; 95% confidence interval [CI], 0.37 – 1.46; *P* = 0.37) (Figure 3). When stratified by age, CCP recipients < 65 years had a 4-fold decrease in mortality (8.8 vs 29.4%; OR, 0.23; 95% CI, 0.05 – 0.95; *P* = 0.04) and a 4-fold decrease in deterioration in oxygenation or mortality (11.8 vs 35.3%; OR, 0.24; 95% CI, 0.06 – 0.87; *P* = 0.03). There was no significant difference in mortality of CCP recipients ≥ 65 years (52.6 vs 45.9%; OR, 1.07; *P* = 0.89) (Figure 3, Table 1). Among the 103 CCP recipients, mortality at day 28 was associated with time from symptom onset to transfusion (OR, 1.12; 95% CI, 1.01 – 1.24; *P* = 0.04), earlier week of admission (OR, 2.22; 95% CI, 1.00 – 5.00; *P* = 0.05) and being Hispanic/LatinX in ethnicity compared to not being Hispanic/LatinX (OR, 8.33; 95% CI, 1.69 – 33.3; *P* = 0.009), adjusted for age, sex, BMI, race, ethnicity, comorbid conditions, week of admission, duration of symptoms, baseline oxygen requirement, corticosteroids, anticoagulation use, D-dimer and lymphocyte counts (Table 2). There was no significant association between CCP NAb or spike protein IgG titers and mortality or oxygenation status in CCP recipients.

**Figure 3.**
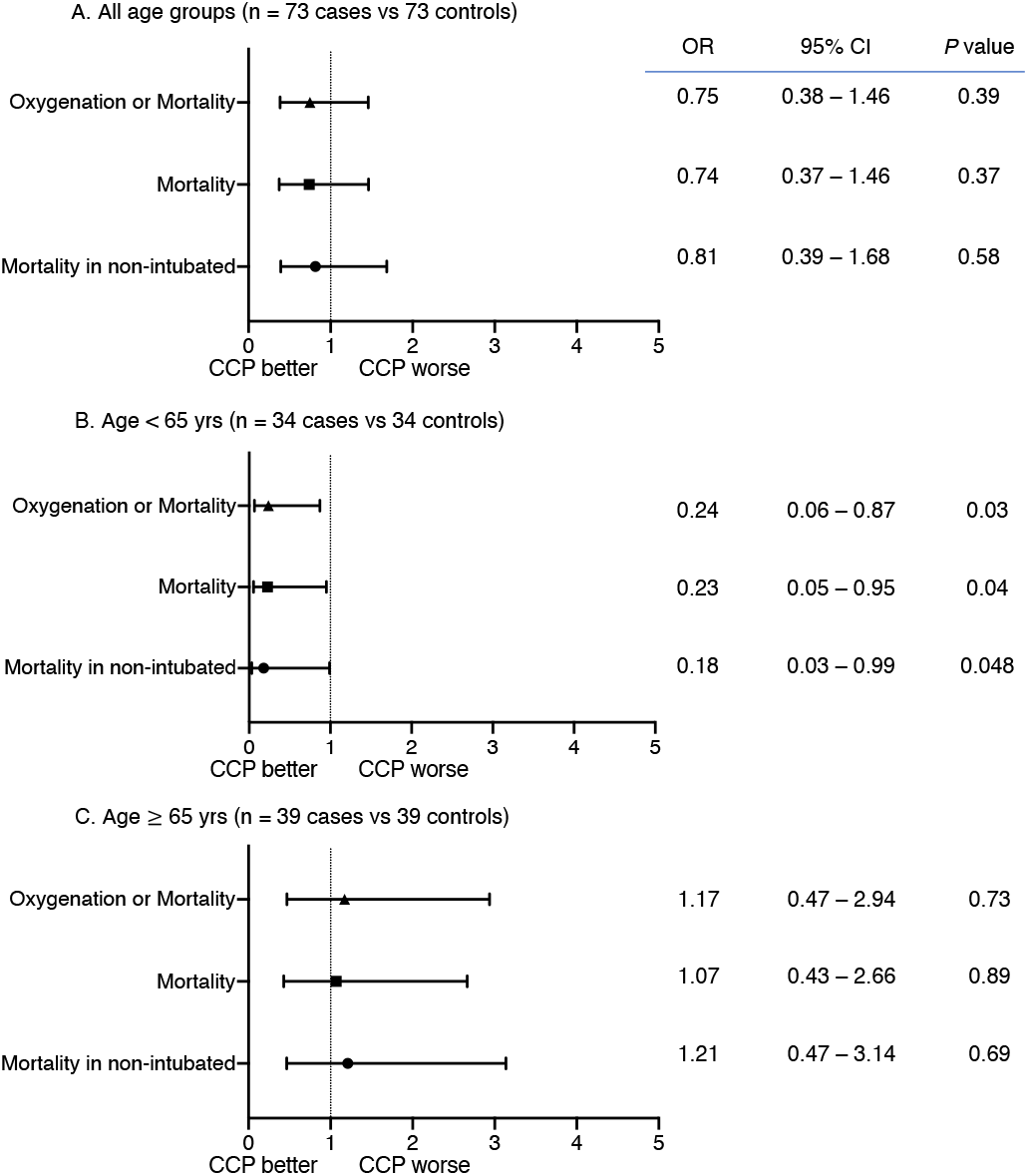
Day 28 outcomes for CCP recipients (n = 73) vs matched controls (n = 73) presented by odds ratio and 95% confidence intervals. A. All age groups (n = 73 cases vs 73 controls). B. Age < 65 years (n = 34 vs 34). C. Age ≥ 65 years (n = 39 vs 39). CCP, COVID-19 convalescent plasma; CI, confidence interval; OR, odds ratio.

**Table 2.**
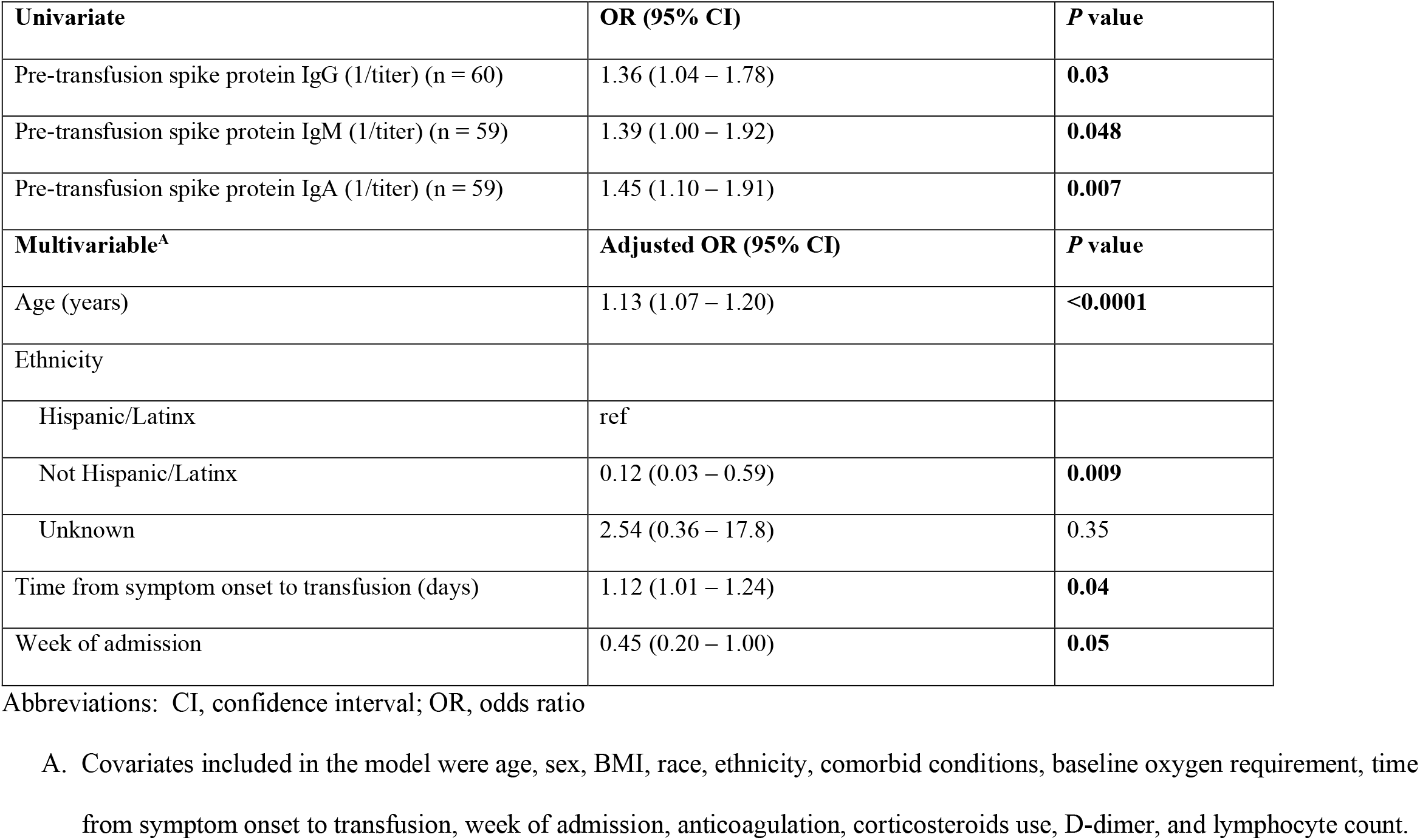
Univariate and Multivariable analysis of mortality at day 28 in all CCP recipients (n = 103).

| **Univariate** | **OR (95% CI)** | **P value** |
| --- | --- | --- |
| Pre-transfusion spike protein IgG (1/titer) (n = 60) | 1.36 (1.04 – 1.78) | **0.03** |
| Pre-transfusion spike protein IgM (1/titer) (n = 59) | 1.39 (1.00 – 1.92) | **0.048** |
| Pre-transfusion spike protein IgA (1/titer) (n = 59) | 1.45 (1.10 – 1.91) | **0.007** |
| **Multivariable<sup>A</sup>** | **Adjusted OR (95% CI)** | **P value** |
| Age (years) | 1.13 (1.07 – 1.20) | **<0.0001** |
| Ethnicity |  |  |
| Hispanic/Latinx | ref |  |
| Not Hispanic/Latinx | 0.12 (0.03 – 0.59) | **0.009** |
| Unknown | 2.54 (0.36 – 17.8) | 0.35 |
| Time from symptom onset to transfusion (days) | 1.12 (1.01 – 1.24) | **0.04** |
| Week of admission | 0.45 (0.20 – 1.00) | **0.05** |

Abbreviations: CI, confidence interval; OR, odds ratio
A. Covariates included in the model were age, sex, BMI, race, ethnicity, comorbid conditions, baseline oxygen requirement, time from symptom onset to transfusion, week of admission, anticoagulation, corticosteroids use, D-dimer, and lymphocyte count.

Multivariable analysis of 90 CCP recipients and 258 controls adjusted for covariates age, sex, BMI, race, ethnicity, comorbid conditions, week of admission, baseline oxygen requirement, corticosteroids, anticoagulation use, D-dimer and lymphocyte counts did not show any difference in outcome between the two groups. When stratified by age, CCP recipients < 65 years had lower mortality or deterioration in oxygenation but this was not statistically significant (OR, 0.23; 95% CI, 0.04 – 1.19; *P* = 0.08) (Supplemental Figure 4). Additionally, multivariable analysis indicated that age, use of mechanical ventilation at baseline, use of systemic corticosteroids, not being on anticoagulation in patients ≥ 65 years and earlier week of admission were associated with mortality at day 28 adjusted for covariates (Supplemental Table 2). Corticosteroid use was associated with mortality in patients receiving low-flow oxygen at baseline (adjusted OR, 2.68; 95% CI, 1.27 – 5.68; *P* = 0.009) when analysis was stratified by baseline oxygen requirement.

### CCP safety and adverse events

There were no adverse reactions, including no instances of transfusion-related acute lung injury or transfusion-associated circulatory overload attributable to CCP administration.

### CCP recipient SARS-CoV-2 spike protein antibody titers

We measured CCP recipient spike protein IgG, IgM, and IgA in remnant serum samples obtained before (D -1) and after transfusion (Figure 4). Baseline spike protein IgG, IgM and IgA were significantly higher in patients mechanically ventilated at enrollment (*P* = 0.009, *P* = 0.01, *P* = 0.02, respectively) and those who died by day 28 (*P* = 0.02, *P* = 0.02, *P* = 0.002, respectively) (Supplemental Table 3, Figure 4, Supplemental Figure 5). There was no association between baseline antibody titers and time from symptom onset to transfusion or time from hospital admission to transfusion. Plateau in median IgG after CCP administration was reached earlier in patients ≥ 65 than < 65 years, mechanically ventilated at baseline versus not, and those who died by versus those alive at day 28 (Figure 4). Baseline IgA was higher in patients ≥ 65 than those < 65 years (*P* = 0.04) (Supplemental Table 3).

**Figure 4.**
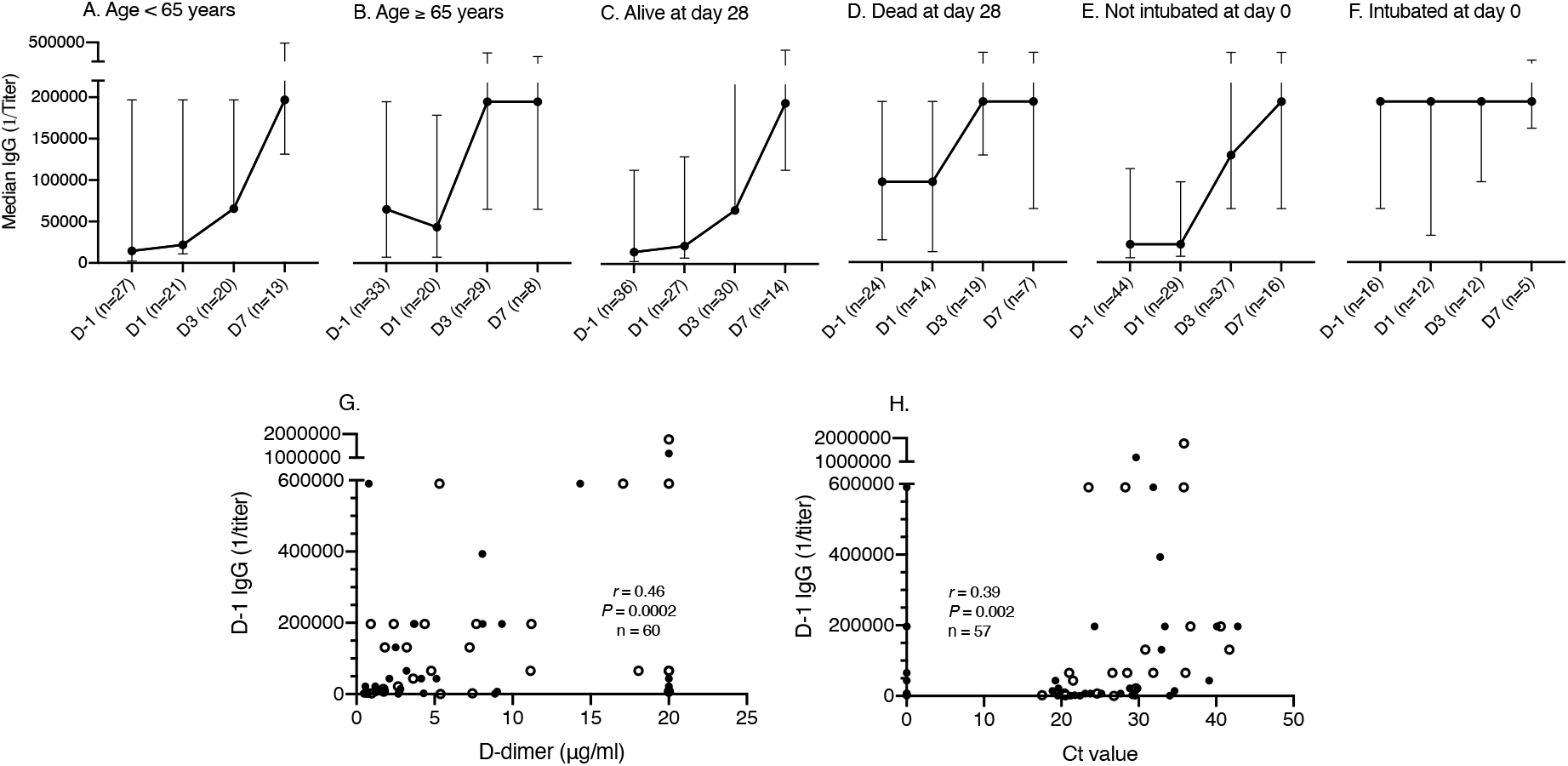
SARS-CoV-2 spike protein IgG titers determined by ELISA at baseline (Day -1 or D-1) and 1, 3 and 7 day after transfusion (D1, D3 and D7) in CCP recipients. (A) Age < 65 years. (B) Age ≥ 65 years. (C) Alive at day 28. (D) Died by day 28. (E) Not intubated on day of transfusion. (F) Intubated on day of transfusion. Correlation between baseline spike protein IgG titer and (G) D-dimer and (H) cycle threshold (Ct) value from initial nasopharyngeal SARS-CoV-2 RT-PCR in CCP recipients. The median titers and interquartile ranges are shown on the y-axis for each time point shown on the x-axis (A-F). X-axis shows days relative to convalescent plasma transfusion (A-F). Open circles show patients who died by day 28 (G, H). r: Spearman’s correlation coefficient. Ct value, cycle threshold value.

There was a direct association between mortality at day 28 and baseline (D -1) IgG (OR, 1.4; 95% CI, 1.04 – 1.78; *P* = 0.03), IgM (OR, 1.39; 95% CI, 1.00 – 1.92; *P* = 0.048) and IgA (OR, 1.45; 95% CI, 1.11 – 1.91; *P* = 0.007) in the univariate analyses but not in the multivariable analysis adjusted for covariates (Table 2). Baseline spike protein IgG titer was significantly correlated with D-dimer (*r* = 0.46; *P* = 0.0002; n = 60) (Figure 4). In addition, there was a direct, albeit weak correlation between baseline spike protein IgG titer and detected viral load measured by cycle threshold (Ct) value of nasopharyngeal SARS-CoV-2 reverse-transcriptase-polymerase-chain-reaction (RT-PCR) (*r* = 0.39; *P* = 0.002; n = 57) (Figure 4). There was no correlation between Ct value and age, duration of illness, or D-dimer.

### CCP recipient inflammatory and hematology measures

There was no significant difference in change in lymphocyte counts, D-dimer, or CRP between day 0 and 28 in CCP recipients compared to controls (data not shown).

## Discussion

CCP has been used as an investigational treatment for COVID-19 since the early days of the pandemic. Numerous observational studies report safety and signals of possible efficacy of CCP in hospitalized patients with COVID-19 (15, 20, 22-25, 35, 36). Here, we report the mortality and clinical outcomes of a cohort of 73 patients with severe to life-threatening COVID-19 who were transfused with one unit of CCP by 72 hours of hospitalization and 73 propensity score matched controls. There was no significant difference in mortality or improvement in oxygenation in CCP recipients compared to controls. Although treated within 72 hours of hospitalization, CCP treated patients had symptom duration of 5-9 days and multiple indicators of severe or life-threatening disease including lymphopenia, elevated D-dimer levels, and the need for supplemental oxygen. In this regard, our findings are similar to those of other studies in which there was no benefit of CCP in hospitalized patients with severe COVID-19 (25, 27, 30). Nonetheless in a subset of patients stratified by age, CCP recipients < 65 years had significantly lower mortality and deterioration in oxygenation by day 28 than controls. Age and duration of symptoms were independently associated with mortality at day 28 in CCP recipients. There were no adverse reactions directly attributable to CCP.

There was no evidence of benefit of CCP in patients ≥ 65 years. Age, a well-documented risk factor for COVID-19 severity and mortality (37, 38), was significantly associated with mortality in unadjusted and adjusted analyses. Patients ≥ 65 years had a higher frequency of co-morbid conditions, higher D-dimer, CRP and SARS-CoV-2 IgA values, and lower lymphocyte levels than those < 65 years, all of which are markers of severe disease (39-44). Notably in published case-control studies in which CCP was associated with reduced mortality, median ages of patients were < 60 years (21, 23, 45), and in the large open label Mayo Clinic study in which there was a signal of reduced mortality in patients who received high titer CCP, 44% of the cohort was < 60, 70% was < 70 years, and CCP was less effective in those > 80 years (22). In addition, a small RCT comparing 80 patients randomized to CCP versus standard of care posted on the pre-print server on November 29^th^, 2020 found a benefit of CCP in patients < 67 years, but not in the entire cohort with a median age of 61 years (46). These data along with ours suggest that ageing may have a detrimental effect on CCP efficacy.

In a case-control study that also did not show a signal of CCP efficacy, Rogers et al. found a higher rate of hospital discharge in patients ≥ 65 years (47). In their study, 14% of CCP recipients were black/African American, 31% were Caucasian/white, 42% were Hispanic/LatinX, and 34% had hypertension, 25% had diabetes, and all received two units of CCP. Our cohort was older, more severely ill, and came from racial/ethnic populations at higher risk for severe COVID-19 and death (48): 26% of CCP recipients were black/African American, 9% were Caucasian/white, 51% were Hispanic/LatinX, which was associated with mortality in our multivariable analysis, and 82% had hypertension, 42% had diabetes, and all received one unit of CCP. This suggests social determinants of health may have adversely affected clinical outcomes of our cohort.

Reflecting practice at the peak of the pandemic at our center, the majority of patients in our cohort received corticosteroids and corticosteroid use was associated with mortality in those requiring low flow oxygen. In another propensity score matched study, corticosteroids use was also associated with higher mortality (49). Among CCP recipients ≥ 65 years in our study, 98% received corticosteroids concurrently with or before CCP. More CCP recipients than controls also received corticosteroids in the Rogers et al. study, which did not find evidence of CCP benefit (47). In addition, although not statistically significant, a higher proportion of CCP recipients than controls received corticosteroids in the Li et al. RCT, in which there was not a signal of CCP efficacy and the median age was 70 years (25). Corticosteroid use has been associated with lower mortality in patients with COVID-19 who require mechanical ventilation (8, 9), and not in the early course of the disease (9, 50). Viral clearance was slower in patients with SARS and MERS who received corticosteroids (51, 52) and corticosteroid use was associated with lower anti-spike protein receptor binding domain (RBD) IgG and neutralization titers in COVID-19 patients (53). Thus, we wonder if corticosteroid use further impaired the immune status of elderly patients in our cohort, who already had lower lymphocyte counts. Data from ongoing RCTs is needed to evaluate the effect of covariates, including corticosteroids on CCP efficacy.

The ability of CCP to affect the course of COVID-19 is most likely a function of viral neutralization early in disease (14, 54). The Mayo Clinic study analysis found that high titer CCP, as defined by signal to cut off ratio on the OrthoV platform, reduced COVID-19 mortality relative to low titer CCP (22). In a propensity score matched study, compared to controls, CCP with a RBD IgG titer ≥1:1,350 reduced mortality in non-intubated patients transfused within 72 hours of hospital admission (21). Although titers in our study cannot be directly compared with titers in other studies, CCP used in our study had high titer spike protein IgG and a median neutralizing titer of 1:938 based, respectively, on a highly specific full length spike protein ELISA (55-57) and a pseudovirus neutralization assay that correlates with live virus (plaque reduction) neutralization (42, 57, 58). Nonetheless, although there was a possible signal of efficacy in the sub-group < 65 years who received CCP within 72 hours of hospitalization, this was not the case in patients ≥ 65 years, who were more severely ill based on baseline data. It is possible elderly patients may require more than one dose of 200 mL of CCP, but two units did not mediate an effect in the Rogers et al. cohort (47). While there are theoretical concerns of antibody-dependent enhancement (ADE) in the presence of sub-neutralizing concentrations of anti-viral antibodies (59, 60), ADE was not reported in other CCP treatment studies and the levels of spike protein antibody with neutralizing capability in CCP in this study make it very unlikely. Given the high mortality of COVID-19 in patients ≥ 65 years and lack of evidence of CCP efficacy in this group in our study and others (25, 61) there is a need for more data on the effect of aging on CCP efficacy in COVID-19.

Consistent with other reports associating SARS-CoV-2 antibody titer with disease severity and/or mortality (53, 62-64), spike protein IgG at enrollment was directly associated with mortality among 60 CCP recipients who had remnant sera available. Although we observed post-transfusion increases in spike protein IgG in patients < 65 years or who were not intubated, this cannot be distinguished from endogenous antibody without comparison to untreated controls. While we did not measure post-transfusion viral loads, pre-transfusion antibody titers correlated with RT-PCR Ct values (inversely correlated with viral load). Thus, as in other studies (53, 65), endogenous antibody may have already contributed to viral control in CCP recipients. The Gharbharan et al. RCT was terminated early when it was discovered majority of the study patients had neutralizing titers at enrollment equivalent to donor CCP (29), although the study appeared on track to meet its expected endpoint. Patients in the PLACID RCT (27) who received CCP had earlier conversion to viral RNA negativity, despite having low levels of neutralizing antibodies at enrollment. In the ConPlas-19 RCT (26), in which 49% of the patients had positive SARS-CoV-2 IgG at enrollment, CCP did not confer benefit, but there was a trend towards reduced mortality. Nonetheless, in our study as in others (27, 66), antibody titers and symptom duration associated with disease severity and a lack of evidence of CCP efficacy. This underscores the longstanding principle that convalescent antibody therapy is most likely to be effective early in the course of viral respiratory diseases (14) as shown for COVID-19 in propensity score matched studies (23, 49) and a recently published outpatient RCT (66).

A strength of our study is that it includes patients over the age of 65 years, who represent a population with disproportionately higher COVID-19 mortality. To date, studies of CCP and other potential therapies for COVID-19 in patients negatively affected by social determinants of health are lacking. The Bronx has a higher poverty rate than the other New York City (NYC) boroughs (5, 67) and our cohort was comprised predominantly of Hispanic or Latinx and black or African American populations who were severely affected by the surge conditions in NYC and experienced higher COVID-19 mortality (68-71). We attempted to control for hospital surge capacity and social determinants of health that can significantly contribute to COVID-19 outcome (72) by including week of admission, race, and ethnicity as propensity scores.

A major limitation of this study is its retrospective and non-randomized study design. The retrospectively identified controls differed in baseline characteristics from the cases, most notably in baseline oxygen requirement, proportion of corticosteroid use and baseline lymphocyte counts, suggesting a mismatch in severity of illness due to selection bias. While propensity score matching and multivariable analysis with adjustments were done to correct for confounding variables, there likely were additional latent and unmeasured variables that were not adjusted for. In addition, time-dependent variables such as hospital bed capacity during surge conditions and advances in clinical practice, such as use of proning techniques, lung protective ventilation strategies, improvement in sedation, may not have been accounted for in our analysis, despite matching and adjusting for baseline week. In addition, poor or absent documentation of oxygen requirement and inconsistencies in obtaining inflammatory markers during the height of the pandemic resulted in missing data, precluding a more complete analysis. Finally, since we could not obtain antibody data for controls, we cannot assess CCP effects on antibody levels.

In summary, we report that CCP administration within 72 hours of hospitalization demonstrated a possible signal of reduced mortality in patients < 65 years. Similar to others, we found CCP was safe with no adverse events directly attributable to transfusion (21, 73, 74). Although our data suggest possible effects of age, disease severity, and corticosteroid use on CCP efficacy, prospective, randomized, controlled trials are needed to establish CCP efficacy and the effect of these and other covariates. Antibody-based therapies, including CCP have now shown promise in outpatients with mild to moderate COVID-19 (66, 75). However, nearly a year into the pandemic, effective therapies for hospitalized patients are still urgently needed, particularly for those who are elderly and at high risk for mortality. If effective in any group of hospitalized patients, CCP will have immense impact on healthcare resources and public health during this ongoing pandemic (76). CCP is rapidly available compared to other pharmaceuticals or vaccines and may be a more feasible option in surge conditions and/or resource limited settings (24).

## Methods

### Patient enrollment

One hundred and three (103) adult patients with laboratory (nasopharyngeal PCR)-confirmed COVID-19 were enrolled in the Mayo Clinic expanded access treatment protocol (31) to receive CCP between April 13 and May 4, 2020. Hospitalized patients were referred to the study team by hospitalists and/or infectious diseases consultants and were deemed eligible to receive CCP if they had been hospitalized for ≤ 3 days or were symptomatic for 3 to 7 days prior to transfusion and had severe and/or life-threatening COVID-19 disease. Patients who were on mechanical ventilation for more than 24 hours were excluded. Severe disease was defined as respiratory symptoms with hypoxemia requiring ≥ 5 liters of nasal cannula oxygen support. Life-threatening disease was defined as respiratory failure requiring mechanical ventilation, shock, and/or multiple organ dysfunction or failure. Ninety-one patients received CCP by day three of hospitalization. Patients or their legally authorized representatives provided informed consent prior to treatment.

### Convalescent plasma procurement and transfusion

After obtaining informed consent, blood was collected between March and April 2020 from otherwise healthy adult volunteers residing in Westchester County, Rockland County and the Bronx, NY who had recovered from COVID-19. Potential donors had a documented positive nasopharyngeal swab by PCR for SARS-CoV-2 during illness and had been asymptomatic for ≥14 days prior to sample collection. Serum was obtained by venipuncture (BD Vacutainer, serum), aliquoted, heat-inactivated at 56°C for 30 minutes and stored at -4°C prior to antibody screening by enzyme-linked immunosorbent assay (ELISA). Donors with SARS-CoV-2 spike protein titers >1:1,000 were referred for apheresis at the New York Blood Center (NYBC). CCP from 46 MMC donors and 8 donors from the general NYBC pool was administered to patients in the study.

Plasma recipients were transfused with 1 unit (∼ 200 mL) of ABO-type matched CCP over 2-3 hours and monitored before, during and after infusion for signs of transfusion-related reactions per standard transfusion protocol.

### Controls and data collection

We identified 1,347 non-CCP recipients with a positive SARS-CoV-2 PCR admitted to MMC between April 13 and May 4 by querying the electronic medical record (EMR). Since most CCP was administered by day 3 post-admission, baseline day was set at day +2 post-admission in non-CCP recipients. Of the 1,347 identified patients, the CCP recipients and 986 non-CCP recipients were excluded because they required < 5 L or no baseline oxygen support, or had a missing baseline oxygen value (n = 903), or were intubated for > 24 hours at baseline (n = 47), or had missing data (n = 36). This resulted in a retrospective control group of 258 non-CCP recipients (Figure 1). We collected age, sex, BMI, race, ethnicity, comorbidities, medications, laboratory findings, and day of death or discharge from CCP-treated and control patients. Additionally, we collected duration of symptoms and hospital day of transfusion from CCP-treated patients from the EMR. Specific laboratory values and clinical characteristics were obtained from EMR by using Structured Query Language.

For both control and CCP recipients, patient oxygen support was evaluated at day 0 and 28 post-CCP and the corresponding day post-admission for controls as follows: low-flow oxygen through nasal cannula or non-rebreather mask (5 - 15 liter), high-flow nasal cannula or non-invasive ventilation, and invasive mechanical ventilation. If a patient’s oxygen requirement increased or the patient died prior to the time-point of interest, patient’s oxygenation status was considered to have worsened. Initial cycle threshold (Ct) value from the SARS-CoV-2 RT-PCR assay performed on a single platform was queried retrospectively for CCP recipients.

### SARS-CoV-2 spike protein IgG, IgM and IgA titers before and after transfusion of CCP

SARS-CoV-2 spike protein-binding IgG, IgM and IgA titers were determined by ELISA using remnant sera obtained from baseline (Day -1 (D-1)) and 1, 3, 7 days after patients received CCP (D1, D3 and D7, respectively). Briefly, microtiter plates (Costar, Corning, NY) were coated with 25 µl of 2 µg/ml purified spike protein (55, 56, 77) in phosphate-buffered saline (PBS) overnight at 4°C, washed with 1X PBS/0.1% Tween (PBS-T), blocked with 3% (v/v) milk (Bio-Rad)/PBS-T for 1 hr at room temperature (RT), washed, and incubated with heat inactivated sera for 2 hrs at RT. Plates were then washed, incubated with isotype-specific horseradish phosphatase (HRP) labeled goat anti-human (GAH) IgG (Thermo Fisher), IgM, or IgA (Millipore Sigma) for 1 hr at RT. Following final washes, plates were incubated with ultra-TMB ELISA substrate (Thermo Scientific) and color development was stopped by addition of 0.5 M sulfuric acid (Sigma Aldrich). Well absorbances at 450 nm (A_450_) were determined using a Cytation 5 (BioTek). The endpoint titer was determined as the highest dilution to give a signal three times the background A_450_ (wells with no sera).

### rVSV-SARS-CoV-2 S Neutralization assay

The neutralization assay was done as previously described (57). Briefly, CCP samples were serially diluted and incubated with pre-titrated amounts of virus for 1 hr at RT, plasma-virus mixtures were added to 96-well plates (Corning) containing monolayers of Vero cells, incubated for 7 hr at 37°C/5% CO2, fixed with 4% paraformaldehyde (Sigma) in PBS, washed with PBS, and stored in PBS containing Hoechst-33342 (1:2,000 dilution; Invitrogen). Viral infectivity was measured by automated enumeration of green fluorescent protein (GFP)-positive cells from captured images using a Cytation 5 automated fluorescence microscope (BioTek) and analyzed using the Gen 5 data analysis software (BioTek). The serum half-maximal inhibitory concentration (NC_50_) was calculated using a nonlinear regression analysis with GraphPad Prism software.

### Study outcomes

The primary outcome was all-cause mortality at day 28 post-CCP. The secondary outcomes were improvement in oxygenation status or mortality at day 28 post-CCP. Exploratory outcomes were associations between pre-CCP SARS-CoV-2 antibody titers and mortality at day 28.

### Statistics

Patient characteristics and outcomes were reported as frequencies and proportions for categorical variables, and median and interquartile range (IQR) for continuous variables. Differences between groups (e.g., CCP versus non-CCP) were determined by student t-test or Mann Whitney U test for continuous variables, and Chi-square or Fischer’s exact tests for categorical variables, as appropriate.

For the outcome analysis, we performed 1:1 propensity score matching using the nearest neighbor matching without replacement on 90 case and 258 control patients to optimize balance of baseline characteristics for assessing the independent effect of CCP on oxygenation and survival. The distribution of O2 requirement prior to matching showed that the cases had higher oxygen requirement (*P* < 0.001). The primary matching criteria included age, sex, race, ethnicity, BMI, week of admission, D-dimer, lymphocyte count, corticosteroids use, anticoagulation use, hypertension, diabetes, chronic pulmonary disease, chronic kidney disease, coronary artery disease, hyperlipidemia with exact matching on baseline oxygen requirement and age group (categorical, < 65 vs ≥ 65 years). Propensity scores were calculated using a logistic regression model. After 1:1 propensity score matching, the analysis included 73 cases and 73 controls and the variables were not significantly different between CCP recipients and controls based on an omnibus test (*P* = 0.80) (78). The all-cause mortality at day 28 post-CCP was depicted by Kaplan-Meier curves. Differences between groups were compared using the log-rank test. Stratification analyses were done by age < 65 vs ≥ 65 years and by baseline oxygen requirement. Corticosteroids and anticoagulation use were not well balanced in the subgroups and were further adjusted for in age-stratified analysis.

As a sensitivity analysis, factors associated with oxygenation status at day 28 were evaluated by proportional odds model, and mortality at day 28 was evaluated using logistic regression model. Adjusted OR and corresponding 95% confidence intervals were calculated. To identify variables that predicted mortality in CCP group, we performed a stepwise model selection using the Akaike Information Criterion (AIC) in a logistic regression model with age, sex, BMI, race, ethnicity, comorbidities, week of admission, time from symptom onset to transfusion, baseline oxygen requirement, anticoagulation, corticosteroid use, D-dimer, and lymphocyte count. Log-transformed SARS-CoV-2 spike protein antibody titers were individually added to select models to evaluate their association with each outcome. A two-sided *P*-value of < 0.05 was considered statistically significant. Prism Version 8.0, Stata/IC Version 16.1 and R was used for analysis.

### Study approval

The retrospective cohort study, the donor plasma procurement protocol and the use of the expanded access protocol was approved by the Albert Einstein College of Medicine Institutional Review Board.

## Data Availability

The data that support the findings of this study are available from the corresponding author upon reasonable request.

## Author contributions

HY, LP, JD, KC designed the study. HY, LP wrote the manuscript with input from all authors. JD, IG, KC, TW contributed to critical revision of the manuscript. HY, RB, KF enrolled patients. MP, LS, JU provided CCP from the blood bank. HY, UNS, KC, SA, JA, AA, DB, AK, BL, AL, ML, AM, XY collected clinical data. IG, RR, RT, PS retrospectively identified control patients and collected clinical data. MRG, ASF provided clinical specimens and critical data interpretation. JR, AN, R Babb collected clinical specimens and performed ELISAs. RHBIII, ASW curated clinical specimens, performed ELISAs, and analyzed ELISA data. EL, CF, DH, MED, JMF assisted with clinical specimen processing and performed ELISAs. DH, MED, RKJ, RHBIII performed and analyzed viral neutralization assays. KT, MCM, OV performed ELISAs. GIG and RJM expressed and purified spike protein. JAQ assisted with management of clinical information. NGH, NCM and SJG expressed, purified and performed quality control on the spike protein. GJK and JMS assisted with specimen collection and transport. RS assisted with specimen collection. DYG provided data. TW, YL, HY performed statistical analysis. HY, LP, JD, KC, TW, IG, UNS, DYG, ASF assisted with data analysis and interpretation. JB acquired funding. All authors revised and approved the final version.

## Acknowledgements

HY was supported by NIH/National Center for Advancing Translational Service (NCATS) Einstein-Montefiore CTSA Grant Number UL1TR001073. LP was supported in part by NIH grants AI 123664, AI 143453, NCATS 3UL1TR002556-04S1 and a grant from the Mathers Foundation. KC was supported in part by NIH grants U19AI142777 and R01AI32633 and a COVID-19 Pilot Award from Albert Einstein College of Medicine. JRL was supported by the NIH (R01-AI125462) and a COVID-19 Pilot Award from Albert Einstein College of Medicine. SCA was supported by R01AI145024, the Wollowick Family Foundation Chair in Multiple Sclerosis and Immunology, and Janet & Martin Spatz and the Helen & Irving Spatz Foundation. Additional support provided by the Albert Einstein Macromolecular Therapeutics Development Facility, the Einstein-Rockefeller-CUNY Center for AIDS Research (P30AI124414), and the Albert Einstein Cancer Center (P30CA013330). JPD was supported by NIAID R21AI141367. MCM and GIG were supported by an NIH Training Program in Cellular and Molecular Biology and Genetics (T32-GM007491). KT, JAQ, RJM, RHBIII and GK were supported by an NIH Medical Scientist Training Program (T32-GM007288) at Albert Einstein College of Medicine. This study was supported in part by a US Department of Health and Human Services (HHS), Biomedical Advanced Research and Development Authority (BARDA) grant contract 75A50120C00096, National Center for Advancing Translational Sciences (NCATS) grant UL1TR002377, Schwab Charitable Fund (Eric E Schmidt, Wendy Schmidt donors), United Health Group, National Basketball Association (NBA), Millennium Pharmaceuticals, Octopharma USA, Inc, and the Mayo Clinic.

## Notes

**Conflicts of Interest statement:** KC is a member of the Scientific Advisory Board of Integrum Scientific, LLC. In addition, KC has a SARS-CoV-2 spike neutralization assay patent pending, and a SARS-CoV-2 spike antibody assay patent pending. JL reports grants from Adimab LLC, grants from Integrated BioTherapeutics, grants from Mapp Biopharmaceutical, personal fees from Johnson & Johnson, personal fees from Celdara Medical. In addition, JL has a COVID-19 antibody diagnostic patent pending. Other authors have declared that no conflict of interest exists.

### Competing Interest Statement

KC is a member of the Scientific Advisory Board of Integrum Scientific, LLC. In addition, KC has a SARS-CoV-2 spike neutralization assay patent pending, and a SARS-CoV-2 spike antibody assay patent pending. JL reports grants from Adimab LLC, grants from Integrated BioTherapeutics, grants from Mapp Biopharmaceutical, personal fees from Johnson & Johnson, personal fees from Celdara Medical. In addition, JL has a COVID-19 antibody diagnostic patent pending. Other authors have declared that no conflict of interest exists.

