## Supplemental material for "Treatment of Severe COVID-19 with Convalescent Plasma in the Bronx, NYC"

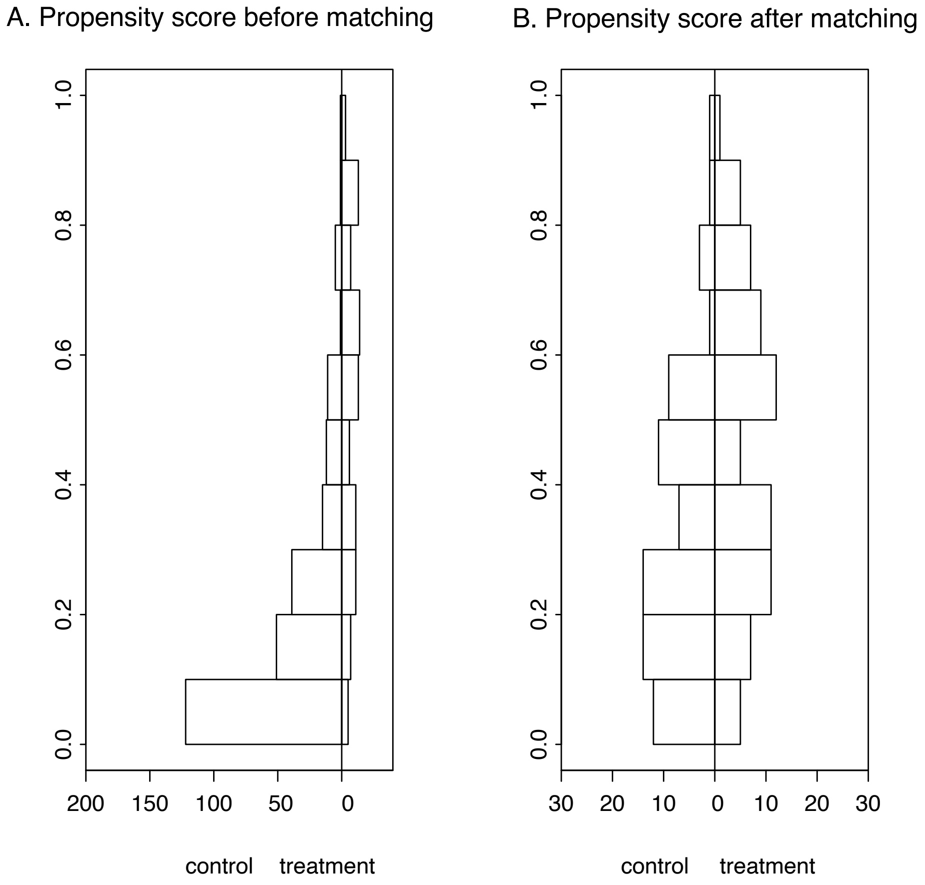

Supplemental Figure 1. Propensity scoring of CCP recipients and matched control patients. All patient comparisons before (A) and after (B) propensity score matching.

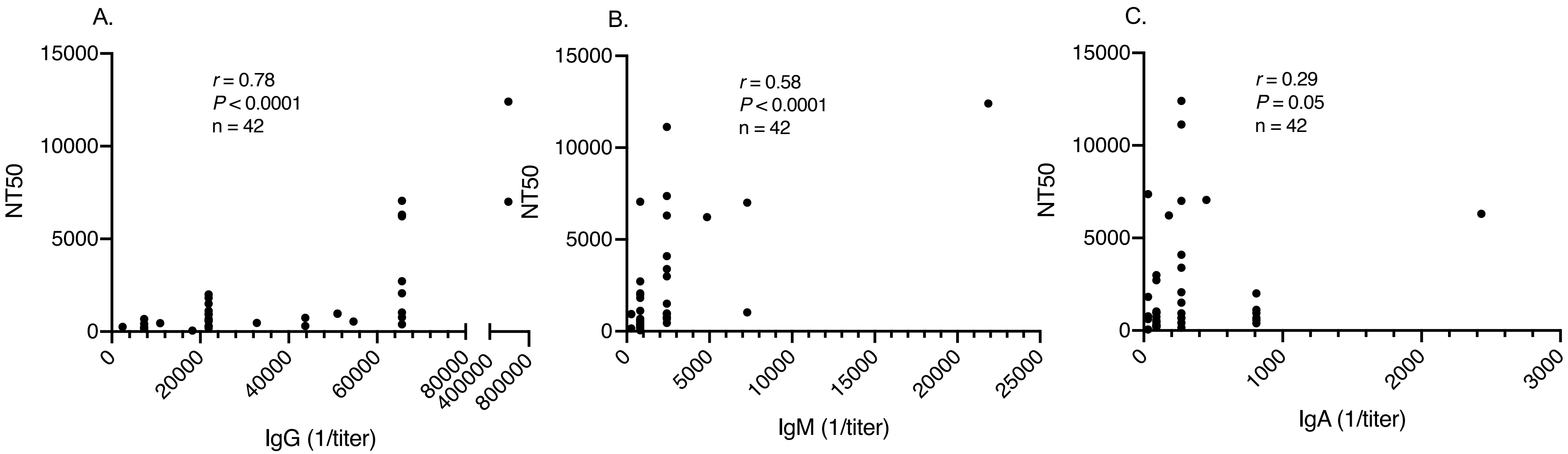

Supplemental Figure 2. Correlation between neutralizing antibody titers (NT50) and spike protein binding (A) IgG, (B) IgM and (C) IgA in CCP determined by ELISA. *r*: Spearman's correlation coefficient.

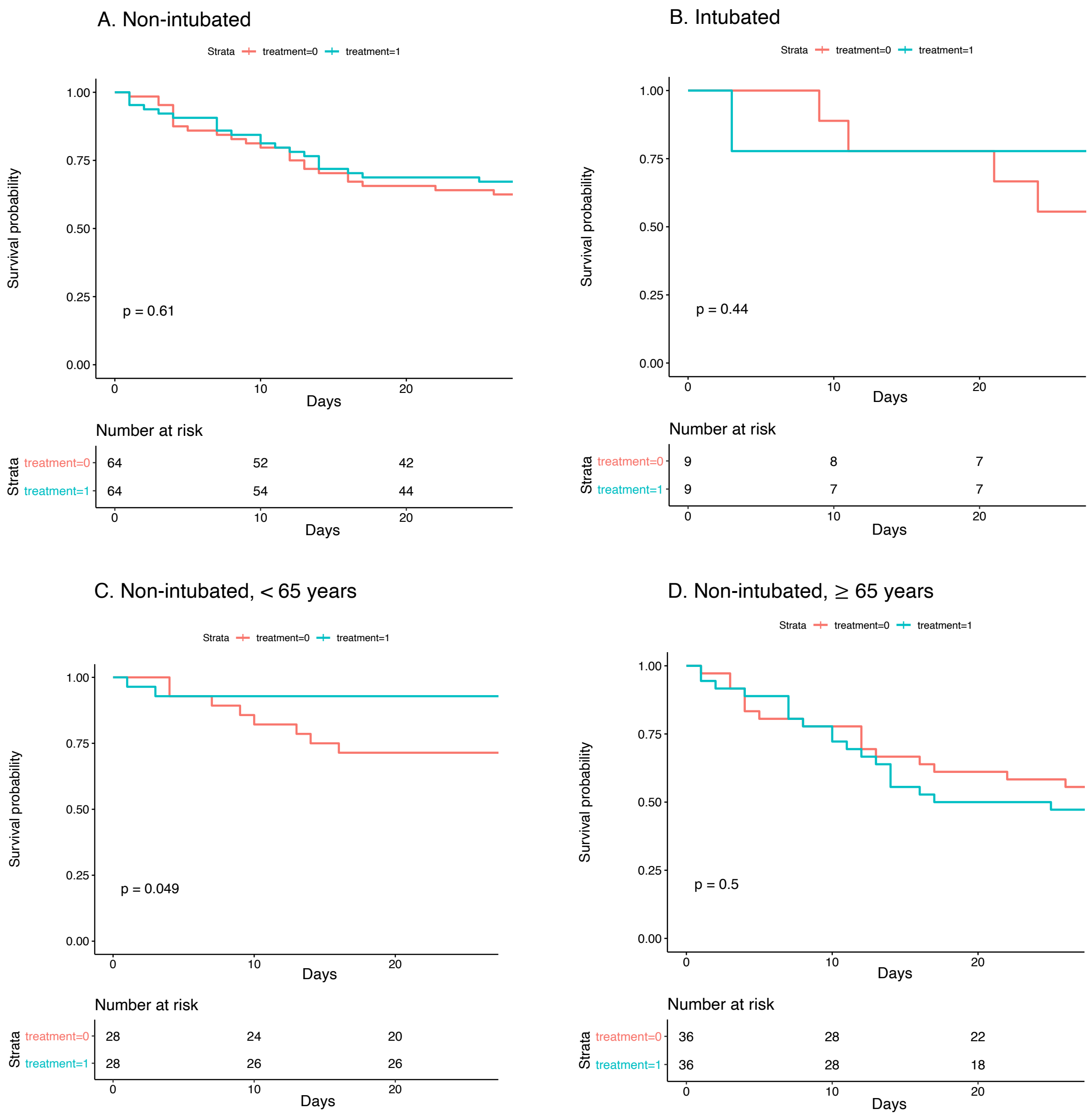

Supplemental Figure 3. Kaplan–Meier Plot of the Probability of Survival from time of transfusion to day 28 in CCP recipients (n = 73) vs matched controls (n = 73) stratified by age and intubation status at baseline. A. Non-intubated. B. Intubated. C. Non-intubated, < 65 years. D. Non-intubated, ≥ 65 years.

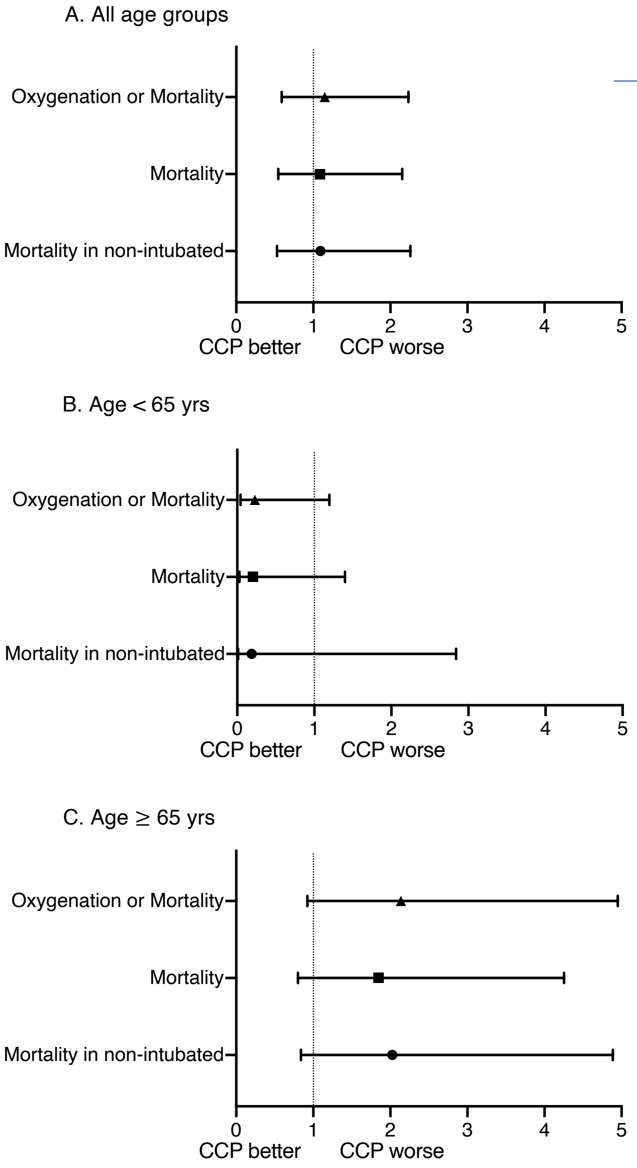

| | OR | 95% CI | *P* value |
|---|---|---|---|
| **A. All age groups** | | | |
| Oxygenation or Mortality | 1.15 | 0.58 – 2.23 | 0.69 |
| Mortality | 1.09 | 0.55 – 2.15 | 0.82 |
| Mortality in non-intubated | 1.09 | 0.53 – 2.26 | 0.81 |
| **B. Age < 65 yrs** | | | |
| Oxygenation or Mortality | 0.23 | 0.04 – 1.19 | 0.08 |
| Mortality | 0.20 | 0.03 – 1.40 | 0.10 |
| Mortality in non-intubated | 0.19 | 0.01 – 2.84 | 0.23 |
| **C. Age ≥ 65 yrs** | | | |
| Oxygenation or Mortality | 2.14 | 0.92 – 4.95 | 0.07 |
| Mortality | 1.85 | 0.80 – 4.25 | 0.15 |
| Mortality in non-intubated | 2.03 | 0.84 – 4.89 | 0.12 |

Supplemental Figure 4. Day 28 outcomes in CCP recipients (n = 90) vs controls (n = 258) presented by odds ratio and 95% confidence intervals in patients. Associations were adjusted for age, sex, BMI, race, ethnicity, comorbidities, baseline oxygen requirements, week of admission, corticosteroids, therapeutic anticoagulation use, D-dimer and lymphocyte counts. A. All age groups. B. Age < 65 years. C. Age ≥ 65 years. CCP, COVID-19 convalescent plasma; CI, confidence interval; OR, odds ratio.

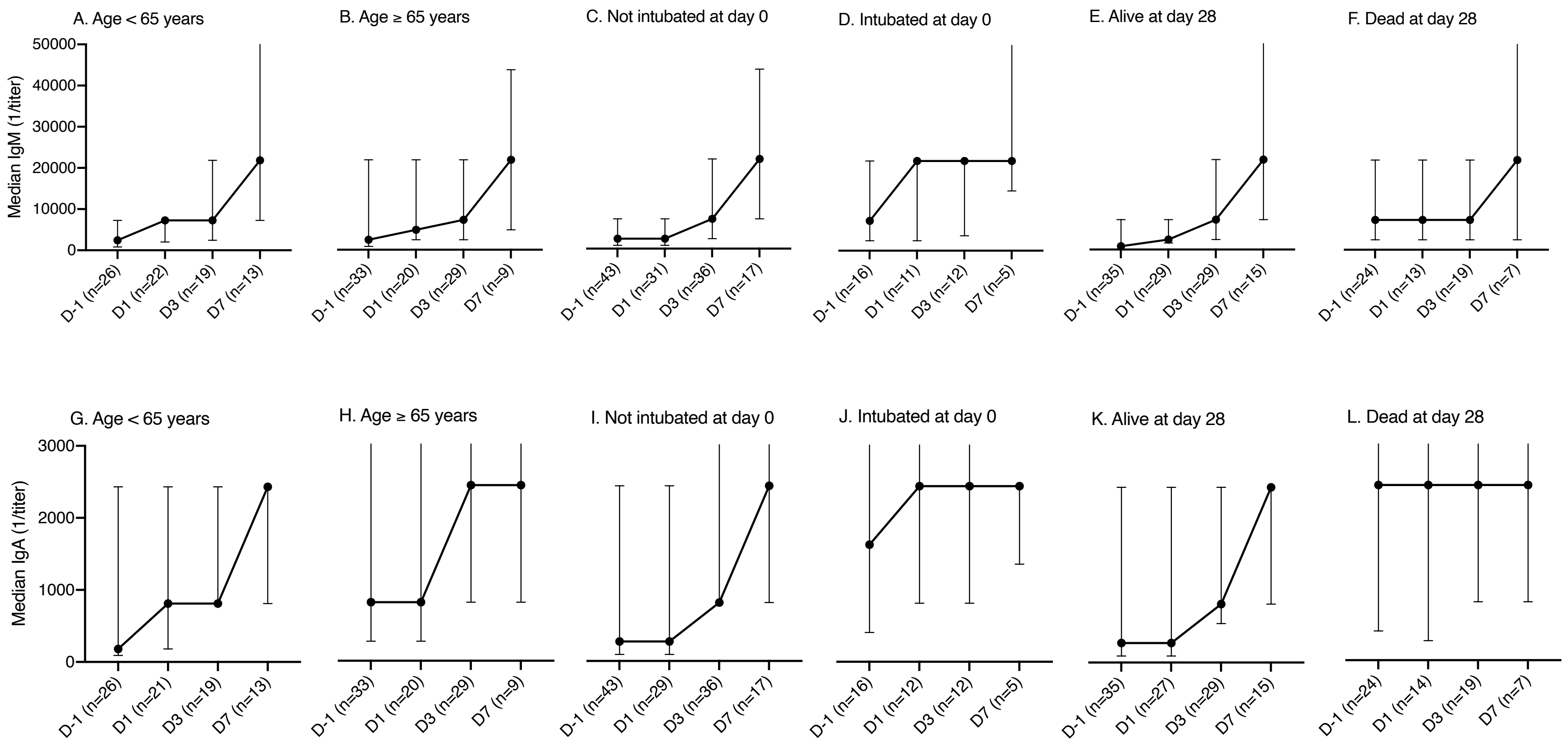

Supplementary Figure 5. SARS-CoV-2 spike protein IgM and IgA titers determined by ELISA at baseline (D-1) and 1, 3 and 7 days after transfusion (D1, D3 and D7) in CCP recipients presented by variables of interest. (A) IgM < 65 years. (B) IgM ≥ 65 years. (C) IgM patients not intubated at day 0. (D) IgM patients intubated at day 0. (E) IgM patients alive at day 28. (F) IgM patients dead at day 28. (G) IgA age < 65 years. (H) IgA age ≥ 65 years. (I) IgA patients not intubated at day 0. (J) IgA patients intubated at day 0. (K) IgA patients alive at day 28. (L) IgA patients dead at day 28. The median titers and interquartile ranges are shown on the y-axis for each time point shown on the x-axis. X-axis shows days relative to convalescent plasma transfusion.

Supplemental Table 1. Baseline characteristics and outcomes in all CCP recipients transfused by admission day 3 (n = 90) and controls before propensity score matching (n = 258).

| | **All patients** | | | **< 65 yrs** | | | **≥ 65 yrs** | | |
|---|---|---|---|---|---|---|---|---|---|
| **Characteristic** | **Case (n=90; 72 not on MV, 18 on MV)** | **Controls (n = 258; 243 not on MV, 15 on MV)** | ***P* value** | **Case (n=43; 36 not on MV, 7 on MV)** | **Controls (n=88; 78 not on MV, 10 on MV)** | ***P* value** | **Case (n=47; 36 not on MV, 11 on MV)** | **Controls (n=170; 165 not on MV, 5 on MV)** | ***P* value** |
| Median age (IQR) - yr | 66 (54-77) | 72 (59-81) | **0.002** | 54 (50-57) | 56 (47.5-59.5) | 0.17 | 77 (70-82) | 78 (72-84) | 0.10 |
| Not on MV | 64.5 (54-75) | 73 (61-81) | **0.001** | 54 (48.5-58) | 56 (48-59) | 0.21 | 75 (70-82) | 78 (72-84) | 0.14 |
| On MV | 67.5 (56-78) | 60 (49-66) | 0.09 | 55 (50-56) | 53.5 (46-60) | 0.84 | 77 (69-79) | 72 (66-75) | 0.21 |
| **Age category – no. (%), yr** | | | | | | | | | |
| <45 | 9 (10.0) | 17 (6.5) | 0.09 | 9 (20.9) | 17 (19.3) | 0.82 | 0 | 0 | |
| 45 to < 65 | 34 (37.7) | 71 (27.5) | | 34 (79.0) | 71 (80.6) | | 0 | 0 | |
| 65 to <75 | 21 (23.3) | 60 (23.2) | | 0 | 0 | | 21 (44.6) | 60 (35.2) | 0.23 |
| 75 to <85 | 20 (22.2) | 70 (27.1) | | 0 | 0 | | 20 (42.5) | 70 (41.1) | |
| ≥85 | 6 (6.6) | 40 (15.5) | | 0 | 0 | | 6 (12.7) | 40 (23.5) | |

| | | | | | | | | | |
|---|---|---|---|---|---|---|---|---|---|
| **Sex, BMI** | | | | | | | | | |
| Male – no. (%) | 51 (56.6) | 152 (58.9) | 0.71 | 27 (62.7) | 57 (64.7) | 0.82 | 24 (51.0) | 95 (55.8) | 0.55 |
| Body-mass index, median (IQR)[A] | 28.3 (23.9-34.5) | 26.7 (22.8-32.2) | **0.05** | 29.3 (27.1-34.9) | 30.3 (25.2-36.1)_ | 0.93 | 26.9 (22.0-32.8) | 25.2 (21.9-29.2) | 0.20 |
| **Ethnicity[B]– no. (%)** | | | | | | | | | |
| Hispanic | 46 (51.1) | 113 (43.8) | 0.26 | 21 (48.8) | 42 (47.7) | 0.45 | 25 (53.1) | 71 (41.7) | 0.39 |
| Non-Hispanic | 32 (35.5) | 117 (45.3) | | 14 (32.5) | 36 (40.9) | | 18 (38.3) | 81 (47.6) | |
| Unknown | 12 (13.3) | 28 (10.8) | | 8 (18.6) | 10 (11.3) | | 4 (8.5) | 18 (10.5) | |
| **Race[B]– no. (%)** | | | | | | | | | |
| White | 8 (8.8) | 32 (12.4) | 0.63 | 3 (6.9) | 6 (6.8) | 0.17 | 5 (10.6) | 26 (15.2) | 0.07 |
| African American | 23 (25.5) | 69 (26.7) | | 16 (37.2) | 20 (22.7) | | 7 (14.8) | 49 (28.8) | |
| Others/Unknown | 59 (65.5) | 157 (60.8) | | 24 (55.8) | 62 (70.4) | | 35 (74.4) | 95 (55.8) | |
| **Coexisting disorder – no. (%)** | | | | | | | | | |
| Hyperlipidemia | 50 (55.5) | 143 (55.4) | 0.98 | 22 (51.1) | 35 (39.7) | 0.21 | 28 (59.5) | 108 (63.5) | 0.62 |
| Hypertension | 74 (82.2) | 222 (86.0) | 0.38 | 30 (69.7) | 65 (73.8) | 0.62 | 44 (93.6) | 157 (92.3) | 0.76 |

| | | | | | | | | | |
|---|---|---|---|---|---|---|---|---|---|
| Coronary artery disease | 14 (15.5) | 32 (12.4) | 0.44 | 3 (6.9) | 8 (9.0) | 1.00 | 11 (23.4) | 24 (14.1) | 0.12 |
| Congestive heart failure | 16 (17.7) | 76 (29.4) | **0.03** | 4 (9.3) | 15 (17.0) | 0.23 | 12 (25.5) | 61 (35.8) | 0.18 |
| Chronic pulmonary disease | 30 (33.3) | 89 (34.5) | 0.84 | 13 (30.2) | 22 (25.0) | 0.52 | 17 (36.1) | 67 (39.4) | 0.68 |
| Chronic kidney disease | 26 (28.8) | 107 (41.4) | **0.03** | 9 (20.9) | 25 (28.4) | 0.35 | 17 (36.1) | 82 (48.2) | 0.14 |
| Obesity (BMI ≥ 30) | 35 (38.8) | 80 (31.0) | 0.17 | 20 (46.5) | 45 (51.1) | 0.61 | 15 (31.9) | 35 (20.5) | 0.10 |
| Morbid obesity (BMI ≥ 35) | 18 (20.0) | 42 (16.2) | 0.42 | 9 (20.9) | 25 (28.4) | 0.35 | 9 (19.1) | 17 (10.0) | 0.08 |
| Diabetes | 38 (42.2) | 121 (46.9) | 0.44 | 14 (32.5) | 33 (37.5) | 0.58 | 24 (51.0) | 88 (51.7) | 0.93 |
| **Symptom duration** | | | | | | | | | |
| Median time from symptom onset to CCP, | 7 (5-9) | n/a | | 8 (5-9) | n/a | | 6 (4-9.5) | n/a | |

| | | | | | | | | | |
|---|---|---|---|---|---|---|---|---|---|
| median days (IQR) | | | | | | | | | |
| **Pharmacologic interventions – no. (%)** | | | | | | | | | |
| Corticosteroids | 84 (93.3) | 163 (63.1) | **<0.0001** | 38 (88.3) | 55 (62.5) | **0.002** | 46 (97.8) | 108 (63.5) | **<0.0001** |
| Therapeutic anticoagulation | 56 (62.2) | 177 (68.6) | 0.26 | 20 (46.5) | 63 (71.5) | **0.005** | 36 (76.6) | 114 (67.0) | 0.21 |
| **Oxygen on the day of transfusion – no. (%)** | | | | | | | | | |
| Low-flow oxygen, 5L - 15L | 61 (67.7) | 208 (80.6) | **<0.0001** | 31 (72.0) | 65 (73.8) | 0.68 | 30 (63.8) | 143 (84.1) | **<0.0001** |
| High-flow oxygen or NIV | 11 (12.2) | 35 (13.5) | | 5 (11.6) | 13 (14.7) | | 6 (12.7) | 22 (12.9) | |
| MV | 18 (20.0) | 15 (5.8) | | 7 (16.2) | 10 (11.3) | | 11 (23.4) | 5 (2.9) | |
| **Laboratory values on the day of transfusion, median (IQR)** | | | | | | | | | |
| Lymphocyte count, $x10^9/L$ | 0.8 (0.5-1.1) | 1.0 (0.8-1.4) | **0.001** | 0.9 (0.7-1.2) | 1.0 (0.8-1.4) | 0.13 | 0.8 (0.5-1.0) | 1.0 (0.7-1.4) | **0.001** |
| Not on MV | 0.8 (0.6-1.1) | 1.0 (0.8-1.5) | **0.001** | 0.9 (0.7-1.1) | 1.0 (0.8-1.5) | 0.07 | 0.8 (0.5-1.0) | 1.0 (0.7-1.4) | **0.002** |
| On MV | 0.8 (0.5-1.6) | 1.1 (0.6-1.2) | 0.82 | 1.2 (0.6-3.9) | 1.1 (0.9-1.2) | 0.55 | 0.6 (0.5-1.6) | 0.8 (0.6-1.2) | 0.73 |

| | | | | | | | | | |
|---|---|---|---|---|---|---|---|---|---|
| D-dimer, μg/ml | 2.7 (1.1-8.3) | 3.3 (1.4-7.9) | 0.20 | 1.2 (0.6-3.2) | 2.3 (1.2-5.6) | **0.01** | 4.3 (2.1-11.2) | 3.5 (1.7-11.0) | 0.28 |
| Not on MV | 2.3 (1-5.4) | 3.3 (1.4-8.5) | **0.02** | 1.1 (0.6-2.7) | 2.3 (1.1-6.0) | **0.003** | 3.8 (2.1-9.9) | 3.5 (1.7-11.0) | 0.61 |
| On MV | 7.8 (2.2-14.3) | 3.6 (1.5-4.7) | 0.13 | 7.6 (1.8-9.3) | 3.1 (1.5-4.7) | 0.52 | 9.7 (2.2-17.0) | 3.8 (2.0-4.2) | 0.28 |
| C-reactive protein, mg/dL | 16.9 (10.3-27) | 15.4 (7.7-27.2) | 0.47 | 14.8 (8.4-22.4) | 14.5 (8.1-26.6) | 0.58 | 18.7 (14.9-28.8) | 15.7 (7.6-27.4) | 0.08 |
| Not on MV | 16.7 (9.5-27.1) | 15.4 (7.8-27.4) | 0.71 | 12.5 (7.3-20.1) | 14.4 (8.2-26.6) | 0.37 | 19.4 (14.8-28.9) | 16 (7.7-27.6) | 0.09 |
| On MV | 17.8 (14.8-24.8) | 12 (6-26.2) | 0.41 | 18.1 (11.5-28) | 18.5 (6.3-26.2) | 0.69 | 16.5 (14.9-22.9) | 9.6 (4.6-12) | 0.22 |
| **Mortality Outcomes – no. (%)** | | | | | | | | | |
| Day 28 mortality | 30 (33.3) | 82 (31.7) | 0.78 | 3 (6.9) | 15 (17.0) | 0.11 | 26 (57.7) | 65 (39.8) | **0.03** |
| Not on MV | 21 (29.1) | 76 (31.2) | 0.73 | 2 (5.5) | 12 (15.3) | 0.13 | 19 (52.7) | 64 (38.7) | 0.12 |
| On MV | 9 (50.0) | 6 (40.0) | 0.56 | 1 (14.2) | 3 (30.0) | 0.45 | 8 (72.7) | 3 (60.0) | 0.61 |
| **Day 28 clinical status – no. (%)** | | | | | | | | | |
| Stable/better | 57 (63.3) | 172 (66.6) | 0.56 | 39 (90.7) | 70 (79.5) | 0.10 | 17 (37.7) | 97 (59.5) | **0.01** |

| | | | | | | | | |
|---|---|---|---|---|---|---|---|---|
| Worse/dead | 33 (36.6) | 86 (33.3) | | 4 (9.3) | 18 (20.4) | | 28 (62.2) | 66 (40.4) |

Abbreviations: BMI, body mass index; IQR, interquartile range; MV, mechanical ventilation; NIV, noninvasive ventilation

A. Body-mass index is the weight in kilograms divided by the square of the height in meters.

B. Information on race and ethnic group was obtained from entries in the medical record, as reported by the patients.

Supplemental Table 2. Multivariable analysis of mortality at day 28 in CCP recipients (n = 90) vs controls (n = 258).

| **Independent Variable** | **Adjusted OR (95% CI)** | ***P* value** |
|---|---|---|
| Treatment | 1.09 (0.55 – 2.15) | 0.82 |
| Age | 1.06 (1.04 – 1.09) | **<0.0001** |
| Female Sex | 0.65 (0.37 – 1.13) | 0.13 |
| Race | | |
| White | ref | |
| Black | 0.73 (0.29 – 1.84) | 0.51 |
| Others | 1.27 (0.49 – 3.29) | 0.62 |
| Ethnicity | | |
| Hispanic | ref | |
| Non-Hispanic | 1.03 (0.46 – 2.34) | 0.93 |
| Others | 0.77 (0.30 – 1.99) | 0.59 |
| BMI | 1.00 (0.96 – 1.04) | 0.97 |
| Admission week | 0.66 (0.48 – 0.92) | **0.01** |
| Baseline O2 requirement | | |
| Low-flow oxygen | ref | |
| High-flow oxygen | 1.69 (0.78 – 3.64) | 0.18 |

| | | |
|---|---|---|
| Mechanical ventilation | 3.29 (1.25 – 8.62) | **0.02** |
| Baseline D-dimer | 1.10 (0.85 – 1.42) | 0.46 |
| Lymphocyte count | 0.75 (0.47 – 1.21) | 0.24 |
| Corticosteroids | 2.38 (1.25 – 4.50) | **0.008** |
| Low-flow oxygen | 2.68 (1.27 – 5.68) | **0.009** |
| < 65 years | 5.08 (0.72 – 35.8) | 0.10 |
| ≥ 65 years | 1.95 (0.94 – 4.04) | 0.07 |
| Therapeutic anticoagulation | 0.63 (0.36 – 1.12) | 0.12 |
| Low-flow oxygen | 0.56 (0.29 – 1.10) | 0.09 |
| < 65 years | 1.49 (0.29 – 7.63) | 0.63 |
| ≥ 65 years | 0.42 (0.21 – 0.83) | **0.01** |
| Hypertension | 0.89 (0.36 – 2.19) | 0.79 |
| Diabetes Mellitus | 0.98 (0.56 – 1.73) | 0.95 |
| Chronic pulmonary disease | 0.77 (0.43 – 1.37) | 0.37 |
| Chronic kidney disease | 1.50 (0.84 – 2.65) | 0.17 |
| Coronary artery disease | 2.56 (1.14 – 5.78) | **0.02** |
| Congestive heart failure | 0.74 (0.38 – 1.45) | 0.38 |
| Hyperlipidemia | 1.47 (0.80 – 2.70) | 0.21 |

Abbreviations:  BMI, body mass index; CI, confidence interval; OR, odds ratio

Supplemental Table 3. Pre-transfusion IgG, IgM and IgA titer by patient variables in CCP recipients.

| Variables | IgG (1/titer), median (IQR) | *P* value | IgM (1/titer), median (IQR) | *P* value | IgA (1/titer), median (IQR) | *P* value |
|---|---|---|---|---|---|---|
| **Age (years)** | | | | | | |
| < 65 | 18,225 (2,430-196,829), n=22 | 0.19 | 2,430 (810-2,430), n=26 | 0.21 | 180 (90-2,430), n=26 | **0.04** |
| ≥ 65 | 54,675 (7,290-196,829), n=38 | | 2,430 (810-21,870), n=33 | | 810 (270-7,290), n=33 | |
| **Sex** | | | | | | |
| Male | 21,870 (2,430-196,829), n=31 | 0.70 | 2,430 (810-21,870), n=30 | 0.72 | 810 (90-3,645), n=30 | 0.69 |
| Female | 43,740 (7,290-196,829), n=29 | | 2,430 (810-7,290), n=29 | | 810 (90-2,430), n=29 | |
| **Body mass index (kg/m$^2$)** | | | | | | |
| < 30 | 43,740 (7,290-196,829), n=42 | 0.82 | 2,430 (810-21,870), n=41 | 0.73 | 810 (90-2,430), n=41 | 0.37 |

| | | | | | | |
|---|---|---|---|---|---|---|
| ≥ 30 | 43,740 (4,253-196,829), n=18 | | 2,430 (810-7,290), n=18 | | 450 (90-2,430), n=18 | |
| **Mechanical ventilation** | | | | | | |
| No | 21,870 (5,468-114,818), n=44 | **0.009** | 2,430 (810-7,290), n=43 | **0.01** | 270 (90-2,430), n=43 | **0.02** |
| Yes | 196,829 (65,610-196,829), n=16 | | 7,290 (2,430-21,870), n=16 | | 1,620 (405-6,075), n=16 | |
| **Hospital day of transfusion** | | | | | | |
| < 3 | 21,870 (7,290-196,829), n=46 | 0.46 | 2,430 (810-7,290), n=45 | 0.38 | 270 (90-2,430), n=45 | 0.41 |
| ≥ 3 | 98,415 (11,138-196,829), n=14 | | 7,290 (810-21,870), n=14 | | 1,620 (210-2,430), n=14 | |
| **Days from symptom onset to transfusion** | | | | | | |
| < 7 | 43,740 (7,290-196,829), n=27 | 0.80 | 2,430 (810-21,870), n=27 | 0.94 | 270 (90-2,430), n=27 | 0.46 |

| | | | | | | |
|---|---|---|---|---|---|---|
| ≥ 7 | 43,740 (7,290-196-829), n=33 | | 2,430 (810-7,290), n=32 | | 810 (90-2,430), n=32 | |
| **Day 28 outcome** | | | | | | |
| Alive | 14,580 (3,038-114,818), n=36 | **0.02** | 810 (810-7,290), n=35 | **0.02** | 270 (90-2,430), n=35 | **0.002** |
| Dead | 196,829 (27,338-196,829), n=24 | | 7,290 (2,430-21,870), n=24 | | 2,430 (405-7,290), n=24 | |

Abbreviations: CCP, COVID-19 convalescent plasma; IQR, interquartile range
